# Self-reported and tracker-estimated physical activity outcomes in women with chronic pelvic pain disorders: A longitudinal evaluation of construct validity

**DOI:** 10.1101/2025.02.10.25322006

**Authors:** Tanisee Nagaldinne, Samia Shahnawaz, Suzanne R Bakken, Noemie Elhadad, Emma N Horan, Carol Ewing-Garber, Jovita Rodrigues, Matteo Danieletto, Kyle Landell, Ipek Ensari

**Affiliations:** Windreich Department of Artificial Intelligence and Human Health, Icahn School of Medicine at Mount Sinai, New York, NY, 10027; Columbia University School of Nursing, New York, NY, 10031; Department of Biomedical Informatics, Columbia University Irving Medical Center, New York, NY, 10031; Department of Biobehavioral Sciences, Teachers College Columbia University, New York, NY, 10027

## Abstract

**Objective:** This study aims to evaluate the short form International Physical Activity Questionnaire (IPAQ) for use in women with chronic pelvic pain disorders (CPPDs) by comparing its scores against objectively-estimated physical activity (PA) outcomes. We investigated IPAQ components that are most consistently predictive of habitual PA behavior.

**Method:** The study sample included 966 weeks of data from 112 women with CPPDs who enrolled in a 14-week mHealth-based self-tracking study. Participants wore Fitbit devices and completed the IPAQ every week. We compared the IPAQ-reported minutes of walking, total activity, sitting, light-, moderate-, and vigorous intensity PA for concordance and divergence against their corresponding Fitbit estimates. We used linear mixed-effects regression models (MLMs) for all analyses and quantified the between-participant variance in the magnitude of agreement between the two methods via random slope terms. We further evaluated temporal consistency in scores using intraclass correlation coefficients (ICCs).

**Results:** IPAQ-reported walking minutes were strongly associated with Fitbit step counts (B = 3952.36; p = 0.006), minutes of moderate PA (B = 15.498; p = 0.0113), and moderate-to-vigorous PA (MVPA; B = 28.973; p = 0.007). IPAQ total activity minutes were associated with Fitbit minutes of vigorous PA (B = 15.183; p = 0.007) and MVPA (B = 25.658; p = 0.010). IPAQ moderate activity minutes were predictive of Fitbit vigorous PA minutes (B = 9.060; SE = 3.719; p = 0.0151). There was substantial between-individual variance in these point estimates based on the significant random-effect terms, and average weekly PA level was a significant moderator of the association between IPAQ-reported and Fitbit-estimated scores for these variables. IPAQ-reported sitting minutes were inversely associated with Fitbit step counts (B = - 3125.61; p = 0.004), and minutes of MVPA (B = -21.848; p = 0.007), vigorous AP (B = -10.854; p = 0.042), and moderate PA (B = -10.985; p = 0.004).

**Conclusion:** These findings provide support for using IPAQ-reported walking and total activity minutes to monitor several PA domains in women with CPPDs, given their concordance with several tracker-estimated PA outcomes. However, the item on “sitting time” may not be a suitable for assessing sedentary time.

## Introduction

Chronic pelvic pain disorders (CPPDs) affect a significant proportion of women worldwide, with prevalence estimates ranging from 5.7-26.6%.^1^ This cluster of conditions, including endometriosis, uterine fibroids, interstitial cystitis/bladder pain syndrome, and other similar diseases characterized by pelvic pain, inflammation, and other related symptoms, significantly impair quality of life (QoL) and well-being.^1–3^ Though scarce, existing studies demonstrate that women with CPPDs engage in less physical activity (PA) and spend more time in sedentary lifestyle.^4,5^ This could be due to numerous reasons, such as worsened physical functioning that hinders PA behavior, CPPD related fatigue, or worry that PA will exacerbate symptoms. Nevertheless, this posits a significant health risk for this population as physical inactivity is considered a major public health risk that is associated with a number of adverse health outcomes including all-cause mortality, cardiovascular and metabolic diseases, obesity, and depression.^3,6,7^ Specifically for those with CPPDs, there is growing recognition of the importance of PA in managing symptoms and improving health outcomes, such as pain reduction, improved mood, and enhanced physical function.^6,8^ Accurate and comprehensive measurement of PA in this population is therefore clinically relevant for monitoring adherence to PA recommendations, assessing intervention efficacy and the relationship of PA to symptom severity, and planning personalized treatments that incorporate appropriate PA goals.

Despite its clinical importance, there is a lack of high-quality data on PA patterns and measurement in CPPDs. Most studies to date rely solely on cross-sectional self-report measures, which are convenient and cost-effective, but also subject to recall bias and potential overestimation of PA levels.^9,10^ Additionally, the accuracy and consistency of self-reported PA estimates may vary depending on the unique characteristics of different patient groups.^11,12^ For example, various chronic disease related factors such as fatigue, sleep quality, or opioid use could impact recall and self-report accuracy.^13^ Despite the increased use of wearables for PA measurement due to their advantages (e.g., low participant burden, accuracy, granularity of data), self-reported measures of PA are still widely used in research and national surveillance systems due to their practicality, low cost, and ease of administration, along with providing contextual information about PA that is not possible to infer from trackers.^14,15^ Systematic reviews conclude that both self-report surveys and consumer grade trackers can meet acceptable accuracy for measuring PA behavior, with variability depending the specific PA domain and user population.^16,17^ This highlights the importance of context and population-specific evaluation of the commonly used standardized PA measures.

The present study aims to address this gap by evaluating the construct validity of the short form International Physical Activity Questionnaire (IPAQ)^18^ among individuals with CPPDs and identify those IPAQ components that are most consistently predictive of habitual PA behavior in this population. The questions on the IPAQ cover domains of walking, moderate- and vigorous-intensity PA, and sedentary time (i.e., weekday sitting minutes) in total weekly minutes. The responses to these questions are multiplied by their respective metabolic equivalent (MET) and a total PA activity score is computed. It is the most widely used self-report measure of PA and has been validated in various populations and settings.^12^ For example, previous research assessing the IPAQ in patients with fibromyalgia and axial spondyloarthritis (axSpA) report varying degrees of weak to moderate agreement between IPAQ- versus Sensewear armband or Actigraph-based estimates of PA (i.e., Pearson’s *r* = 0.04-0.19 for fibromyalgia; *r* = 0.032-0.367 for axSpA).^19,20^ Another study conducted with healthy adults^21^ reported small (sedentary, moderate, and walking minutes, *r* = 0.16, 0.09, and 0.17, respectively, *p* < 0.001) to moderate (vigorous intensity and total PA; *r* = 0.29 and 0.40, *p* < 0.001) correlations between synchronous paired measurements from Fitbits and the IPAQ. However, IPAQ has not been comprehensively evaluated for use among patients with CPPDs. This is an important point of inquiry because the nature of the disease might differentially impact the degree to which the IPAQ domains are predictive of their reference benchmark (e.g., tracker estimates). Elucidating these potential differences is necessary for making informed decisions on how to best measure PA behavior in this population.

Accordingly, the objectives of this study are threefold: 1) Evaluate construct validity through concordance and divergence between IPAQ-reported vs Fitbit-estimated scores on different PA domains, 2) Examine between-individual variability in the agreement between the two measurement methods, and 3) Assess temporal consistency between the methods through comparison of the magnitude of variability over time. From a measurement science perspective, this evaluation requires consideration of the dynamic nature of PA behavior and any systematic individual differences in measurement.^22^ We address these aspects through a longitudinal, repeated-measures design and a flexible mixed-effects multilevel modeling (MLM) framework. We hypothesized that: 1) IPAQ-reported minutes of walking and different intensities of PA would positively correlate with Fitbit-estimated step counts and moderate- and vigorous intensity PA scores, and negatively correlate with Fitbit-estimated sedentary minutes, 2) IPAQ-reported sitting minutes would positively correlate with Fitbit-estimated sedentary minutes, and negatively correlate with Fitbit-estimated minutes of all PA intensities, and 3) there would be substantial variability in the between-individual variance in the point estimates and a moderating effect of habitual PA levels on the association of IPAQ-reported to Fitbit-estimated outcomes.

## Methods

### Study Design and Data Collection

The study design and procedures were approved by the IRB of the Icahn School of Medicine at Mount Sinai (ISMMS; IRB# STUDY-22-01002). This is a secondary analysis of the data from an ongoing larger study that aims to design, develop, and evaluate CPPD-specific mHealth measures from patient generated health data with high complexity and temporality using non-linear distributed lag and functional data modeling (NIH/NICHD: R01HD108263). It uses an observational study design to collect 90 days of data on patient self-tracked symptoms via a research mHealth app *ehive*^23^ and passively collected activity data using activity trackers from participants. All participants used the ehive research study app for providing the baseline and weekly data on overall health, symptoms, well-being and health behaviors, as well as for receiving prompts and reminders about the study.^23^ Participants were instructed to wear a Fitbit for the duration of the study on their non-dominant wrist continuously for the 14-week study period, except during water-based activities and charging. They also completed the IPAQ short form at the end of each week, reporting their PA behavior for the preceding 7 days.

### Study Sample

Recruitment and enrollment procedures are explained in detail elsewhere.^24^ Briefly, participants were recruited from all campuses of the Mount Sinai Health System (MSHS) and Columbia University Irving Medical Center (CUIMC) via email advertisements and on the myChart by EPIC mobile app for MSHS patients. Inclusion criteria were: (1) individuals with a female reproductive anatomy and diagnosis of a CPPD (e.g., endometriosis, uterine fibroids, interstitial cystitis, or chronic pelvic pain syndrome), (2) experiencing pelvic pain for at least 6 months, and (3) ability to read and understand English. Exclusion criteria included pregnancies and any major comorbidities. All participants provided written informed consent.

### Data Processing

Raw Fitbit data were downloaded and aggregated into weekly totals for steps, sedentary minutes, and minutes of light, moderate, and vigorous physical activity. IPAQ responses were scored according to the IPAQ scoring protocol^25^ to derive weekly totals for minutes of walking, and moderate- and vigorous intensity PA. For data quality, weeks with fewer than 4 valid days of Fitbit wear time (≥10 hours/day) were excluded from the analysis.^26^ IPAQ and Fitbit data were cleaned according to standard protocols, removing outliers and implausible values. Weekly MVPA for the Fitbit and IPAQ were calculated by adding up the weekly totals of moderate-intensity activity and vigorous-intensity activity. Total weekly activity for the IPAQ was calculated by adding up minutes of walking, moderate-, and vigorous-intensity PA.^27^

### Data Analysis

We calculated descriptive statistics for all outcomes and demographics to characterize the study sample. While bivariate correlational analyses and metrics are the traditionally used for measure evaluation, they are not suitable for nested data where there is a distinct grouping structure and when the outcome isn’t expected to be time-invariant.^28^ We instead used MLMs^29^ to compare the IPAQ-based and Fitbit-estimated scores on multiple PA domains for concordance and divergence, and the between-individual variance in these associations. We used intraclass correlations (ICCs) within a MLM framework to evaluate variability in scores and consistency in inter-method agreement, as this framework accommodates the nested structure of the data

#### Temporal Consistency and Intraclass Correlations

Given the non-time-invariant nature of PA behavior, we assessed agreement across the 14 weeks through comparison of the amount of variability in scores captured by the two methods. ICCs are well-suited for this purpose as they can quantify the degree of absolute agreement among measurements and account for systematic differences between measurement methods.^30^ First, we conducted MLMs with bootstrapping for significance testing^31^ to estimate within- and between group variance with 95% confidence intervals (CIs). The resulting “repeatability” statistic is analogous to the ICC^31,32^ where higher R values indicate stronger group-level effects (i.e., between-group variance) and therefore more consistent within-group measurements. Published recommendations categorize the magnitude of the values as weak (0.0 - 0.3), moderate (0.3 - 0.7), and strong (0.7 - 1.0).^29^

Next, we calculated inter-rater reliability (ICC2r) and intra-rater reliability (ICC2a) via two-way random effects model (i.e., ICC(2,1))^33^ using the R irrICC library.^34^ Relying on the methodology by Gwet,^34,35^ they are computed using variance decomposition (i.e., σ²subject, σ²rater, σ²error) where; ICC2r = σ²s/(σ²s + σ²r + σ²e) and ICC2a = (σ²s + σ²r)/(σ²s + σ²r + σ²e). ICC2r represents absolute agreement between methods (i.e., “raters”), while ICC2a represents the pooled intra-method (i.e., “rater”) consistency over time. The key difference is that ICC2a accounts for systematic differences between the two methods by including the rater variance (σ²r) in the numerator. To assess how individual response patterns affect the agreement between the two methods (i.e., generalizability across the sample), we computed ICC2a in models with a subject-rater interaction (i.e., σ²sr).^36^ The no-interaction model assumes a consistent relationship between IPAQ and Fitbit measurements across participants, while the interaction model accounts for participant-specific patterns in how the two methods relate.^34,36^

#### Concordance and Divergence Analyses

To assess construct validity, we evaluated concordance and divergence between IPAQ- vs Fitbit-based PA scores. We conducted separate MLMs for each IPAQ domain (i.e., steps/walking, minutes of PA intensities, total PA minutes, sedentary/sitting) to evaluate their concordance and divergence with the Fitbit-estimated PA variables. For concordance (i.e., expected positive association) evaluation, we regressed the Fitbit-estimated outcome on its corresponding IPAQ-based outcome as the primary predictor. For divergence evaluation (i.e., expected inverse association), we regressed Fitbit-estimated PA scores on the IPAQ-based sitting minutes scores as the predictor. For all models, we scaled the IPAQ regressor (i.e., by subtracting their mean and dividing by their standard deviations) in the models as this is standard and recommended for better convergence. We further included the number of weeks in the study as a covariate to adjust for the number of measurements per participant.

All models included a random intercept term for participant to account for the hierarchical data structure and correlated errors. The random intercept estimate allows comparison of the proportion of variance in the PA outcome attributable to between-participant differences. Based on our a priori hypothesis that the fixed effects would vary by participant, we also estimated a random slope term for the IPAQ-based predictor in each model based on recommendations for fitting MLMs.^37^ This allows the slope of the point estimate to vary across participants and evaluates the between-individual variance in the predictor point estimate. The model estimated correlation between the random intercept and slope indicates the relationship between the intercepts and coefficients. A statistically discernable random slope (p<0.05 based on the single-term deletion likelihood ratio test statistic; LRT) indicates significant between-individual variance in the predictor point estimate. If the variance in the random slopes is estimated close to, or at, zero, the model containing random slopes will have a singular fit and therefore convergence error. In these instances, the random slope component was removed from the final model, as per guidelines.^38^

#### Moderator Analysis

We assessed habitual MVPA level as a potential moderator of the relationship between self-reported IPAQ and Fitbit-estimated outcomes. In each MLM, we included an interaction term between each participant’s habitual weekly PA (i.e., weekly MVPA minutes averaged across their total number of weeks) and the IPAQ predictor variable, in addition to the random intercept and total number of weeks as co-variate as before. All analyses were performed using R version 4.0.3, with the lme4, lmerTest, and rptR libraries in R/RStudio. Statistical significance was set at p < 0.05.

## Results

### Study Sample Characteristics

Summary statistics for the study sample demographics and PA outcomes are provided in Table 1. The final analytic sample included 966 person-level weeks of data from 112 women aged 18-54 years (mean age 35.59 ± 8.70 years). On average, participants provided 10.81 ± 3.36 weeks of data each. The majority of participants (65.28%) had a diagnosis of endometriosis, 16.67% had uterine fibroids, and 27.78% had other forms of chronic pelvic pain disorder (Table 1). Body Mass Index (BMI) of the sample ranged from 15.96 to 48.14 kg/m² (mean 26.42 ± 5.91 kg/m²). The highest level of education was a college degree or higher (84.7%).

**Table 1.**
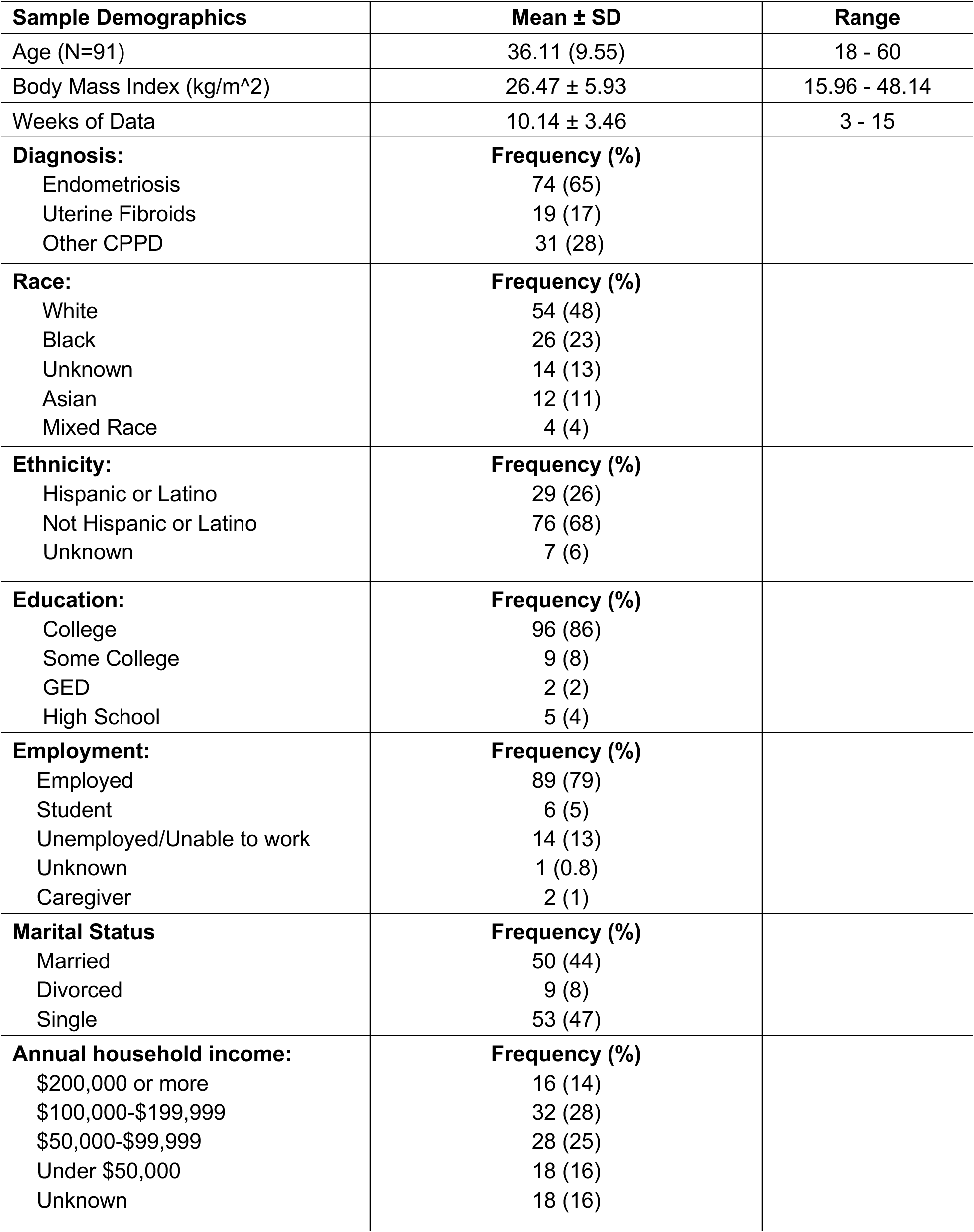
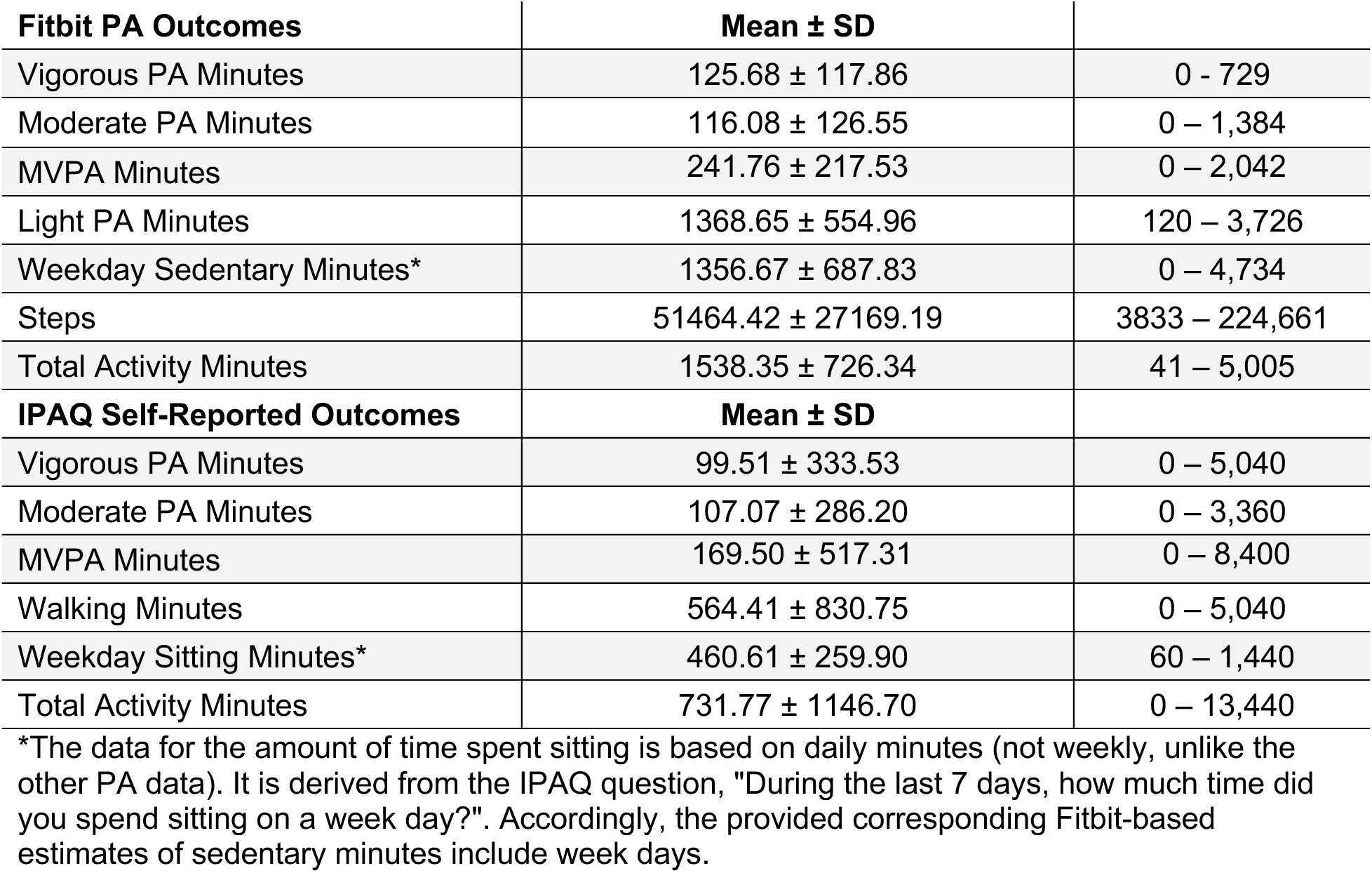
Study Sample Characteristics.

| **Sample Demographics** | **Mean ± SD** | **Range** |
| --- | --- | --- |
| Age (N=91) | 36.11 (9.55) | 18 - 60 |
| Body Mass Index (kg/m<sup>2</sup>) | 26.47 ± 5.93 | 15.96 - 48.14 |
| Weeks of Data | 10.14 ± 3.46 | 3 - 15 |
| **Diagnosis:** | **Frequency (%)** |  |
| Endometriosis | 74 (65) |  |
| Uterine Fibroids | 19 (17) |  |
| Other CPPD | 31 (28) |  |
| **Race:** | **Frequency (%)** |  |
| White | 54 (48) |  |
| Black | 26 (23) |  |
| Unknown | 14 (13) |  |
| Asian | 12 (11) |  |
| Mixed Race | 4 (4) |  |
| **Ethnicity:** | **Frequency (%)** |  |
| Hispanic or Latino | 29 (26) |  |
| Not Hispanic or Latino | 76 (68) |  |
| Unknown | 7 (6) |  |
| **Education:** | **Frequency (%)** |  |
| College | 96 (86) |  |
| Some College | 9 (8) |  |
| GED | 2 (2) |  |
| High School | 5 (4) |  |
| **Employment:** | **Frequency (%)** |  |
| Employed | 89 (79) |  |
| Student | 6 (5) |  |
| Unemployed/Unable to work | 14 (13) |  |
| Unknown | 1 (0.8) |  |
| Caregiver | 2 (1) |  |
| **Marital Status** | **Frequency (%)** |  |
| Married | 50 (44) |  |
| Divorced | 9 (8) |  |
| Single | 53 (47) |  |
| **Annual household income:** | **Frequency (%)** |  |
| \$200,000 or more | 16 (14) | |
| \$100,000-\$199,999 | 32 (28) | |
| \$50,000-\$99,999 | 28 (25) | |
| Under \$50,000 | 18 (16) | |
| Unknown | 18 (16) |  |

| **Fitbit PA Outcomes** | **Mean ± SD** |  |
| --- | --- | --- |
| Vigorous PA Minutes | 125.68 ± 117.86 | 0 - 729 |
| Moderate PA Minutes | 116.08 ± 126.55 | 0 – 1,384 |
| MVPA Minutes | 241.76 ± 217.53 | 0 – 2,042 |
| Light PA Minutes | 1368.65 ± 554.96 | 120 – 3,726 |
| Weekday Sedentary Minutes* | 1356.67 ± 687.83 | 0 – 4,734 |
| Steps | 51464.42 ± 27169.19 | 3833 – 224,661 |
| Total Activity Minutes | 1538.35 ± 726.34 | 41 – 5,005 |
| **IPAQ Self-Reported Outcomes** | **Mean ± SD** |  |
| Vigorous PA Minutes | 99.51 ± 333.53 | 0 – 5,040 |
| Moderate PA Minutes | 107.07 ± 286.20 | 0 – 3,360 |
| MVPA Minutes | 169.50 ± 517.31 | 0 – 8,400 |
| Walking Minutes | 564.41 ± 830.75 | 0 – 5,040 |
| Weekday Sitting Minutes* | 460.61 ± 259.90 | 60 – 1,440 |
| Total Activity Minutes | 731.77 ± 1146.70 | 0 – 13,440 |

\*The data for the amount of time spent sitting is based on daily minutes (not weekly, unlike the other PA data). It is derived from the IPAQ question, "During the last 7 days, how much time did you spend sitting on a week day?". Accordingly, the provided corresponding Fitbit-based estimates of sedentary minutes include week days.

### Temporal Consistency and Intraclass Correlations

Results of the MLMs comparing temporal consistency for all PA domain scores are provided in Table 2. For the IPAQ, the highest repeatability was observed for walking minutes (R = 0.766), followed by total PA Minutes (R = 0.642) and sitting minutes (R = 0.595). For Fitbit-estimated outcomes, step counts had the highest consistency (R = 0.601), followed closely by moderate PA minutes (R = 0.596) and vigorous PA Minutes (R = 0.561). The repeatability values for the remaining PA domains were lower, though statistically significant. The results of the inter- and intra-method reliability analyses are provided in Table 2 (Columns 5,6,7). Walking minutes/steps demonstrated the strongest agreement (i.e., 0.402 for both ICC2r and ICC2a), indicating more consistent absolute agreement between methods. Total PA was associated with lower inter-method agreement (i.e., ICC2r = 0.202) and higher overall intra-method consistency (ICC2a = 0.555). Sedentary behavior had the largest difference between the 2 agreement types (i.e., ICC2r = 0.101 vs ICC2a = 0.650). Models accounting for subject-method interaction yielded increased ICCa values for all PA domains, with the largest change observed for walking mins/step counts.

**Table 2.** Results from repeatability (R) and Intraclass correlation (ICC) estimation. ICC2r and ICC2a values denote inter- and intra-method reliability without subject-method interaction.

| **IPAQ Variable**<br>(N*, n**) | **R (SE)** | **Fitbit Variable**<br>(N=966, n=112) | **R (SE)** | **ICC2r** | **ICC2a** | **ICC2a w/**<br>**interaction** |
| --- | --- | --- | --- | --- | --- | --- |
| Walking Min<br>(N=602, n=102) | 0.766 (0.031),<br>95% CI = 0.697, 0.817 | Step Counts | 0.601 (0.038)<br>95% CI= 0.513, 0.666 | 0.402 | 0.402 | 0.656 |
| Total PA Min<br>(N=423, n=88) | 0.642 (0.045),<br>95% CI = 0.542, 0.721 | Total PA Min<br>(N=926, n=109) | 0.470 (0.042)<br>95% CI = 0.384, 0.547 | 0.202 | 0.555 | 0.769 |
| MVPA Min<br>(N=539, n=100) | 0.154 (0.043),<br>95% CI= 0.070, | MVPA Min | 0.578 (0.038)<br>95% CI = 0.497, 0.645 | 0.152 | 0.171 | 0.288 |
| Vigorous PA Min<br>(N=803, n=112) | 0.250 (0.040),<br>95% CI = 0.171, 0.326 | Vigorous PA<br>Min | 0.561 (0.038)<br>95%CI = 0.479, 0.630 | 0.147 | 0.151 | 0.309 |
| Moderate PA Min<br>(N=748, n=109) | 0.224 (0.042)<br>95% CI = 0.142, 0.308 | Moderate PA<br>Min | 0.596 (0.037)<br>95% CI = 0.518, 0.665 | 0.152 | 0.152 | 0.305 |
| Sitting Min<br>(N=540, n=90) | 0.595 (0.050),<br>95% CI = 0.486, 0.686 | Week day<br>Sedentary Min | 0.303 (0.040)<br>95% CI= 0.225, 0.383 | 0.101 | 0.650 | 0.688 |
|  |  | Light PA Min | 0.547 (0.039)<br>95% CI = 0.466, 0.621 |  |  |  |

\*Number of person-level weeks of data included in the model, \*\*Number of participants MVPA=moderate-to-vigorous intensity physical activity. 95% CI= 95% confidence intervals

### Concordance and Divergence Analyses

The results from the MLMs evaluating concordance of IPAQ scores to FitBit estimates are provided in Table 3. IPAQ-reported walking minutes showed significant positive associations with several Fitbit-measured outcomes, including step counts (B = 10750.15, p = 0.0001), light PA minutes (B = 66.06, p = 0.0124), moderate PA minutes (B = 36.371, p = 0.0026), and MVPA minutes (B = 78.721, p = 0.0014). Reverse-scaling for easier interpretation, these point estimates suggest that for every 1-minute increase in IPAQ-reported walking; Fitbit step counts increase by approximately 8.93 steps, Fitbit light PA minutes increase by 0.055 minutes, Fitbit moderate PA minutes increase by 0.030 minutes, and Fitbit MVPA minutes increase by 0.065 minutes. IPAQ-reported total activity minutes were significantly associated with Fitbit vigorous PA minutes (B = 29.532, p = 0.0106) and Fitbit MVPA minutes (B = 57.319, p = 0.0095). When reverse-scaled, these correspond to increases in Fitbit MVPA by 0.066 minutes and in Fitbit vigorous PA minutes by 0.034 minutes, for every 1-minute increase in IPAQ-reported total activity. The models evaluating the association of IPAQ sitting minutes to Fitbit-estimated sedentary minutes were not statistically significant (p>0.05). Finally, all random slope terms were positive and statistically significant (See Figures 1-5), indicating that individuals with higher baseline levels of a given PA outcome (i.e., intercepts) also tend to have steeper increases in the Fitbit-estimated scores as their IPAQ-reported PA increase (i.e., slopes).

**Figure 1.**
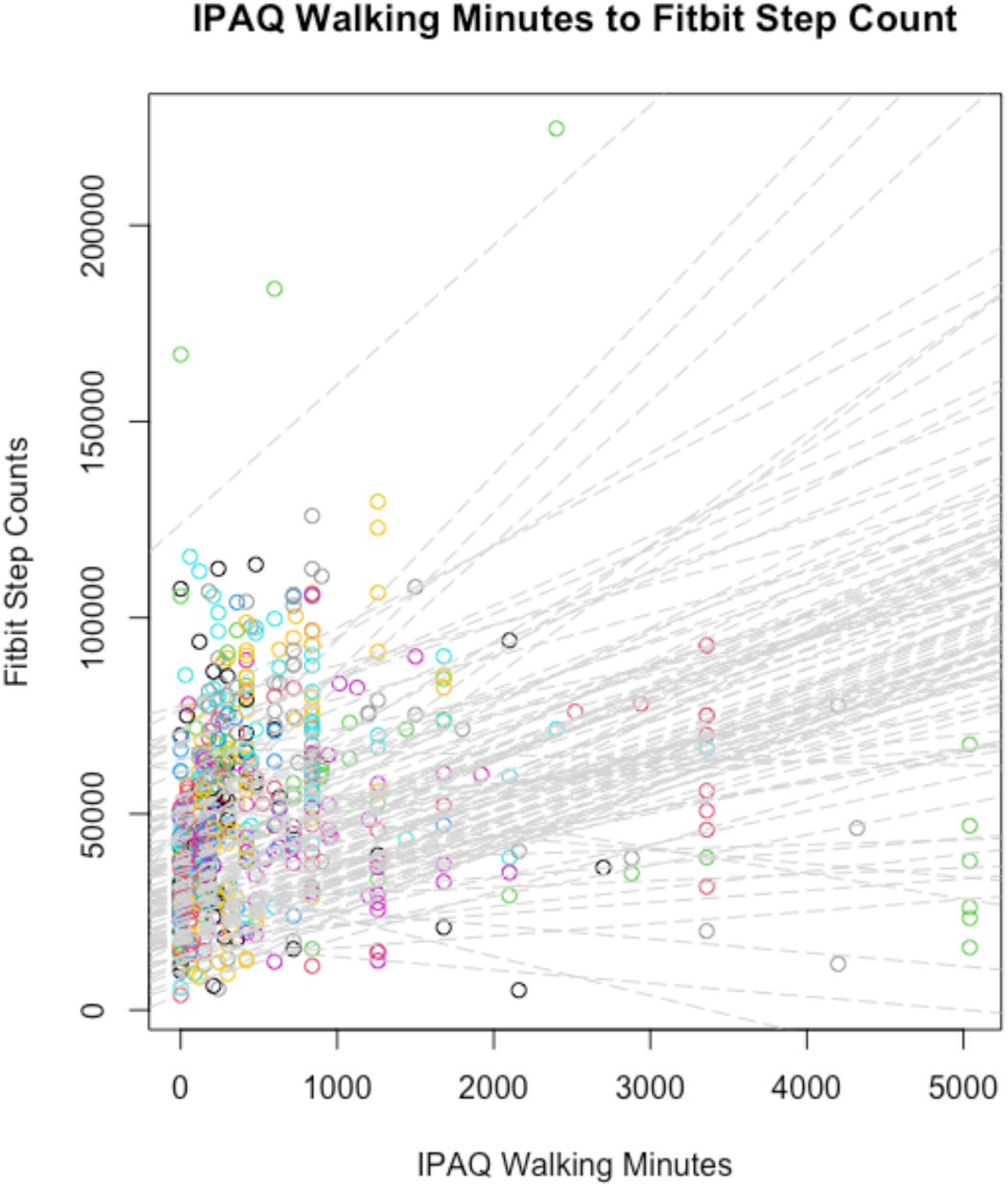
Model-estimated random (i.e., person-level) slopes for the model predicting Fitbit step counts (y-axis) from IPAQ-based walking minutes. Each participant is represented by one dotted grey line (n=102). Each colored circle represents one person-level week (N=602).

**Figure 2.**
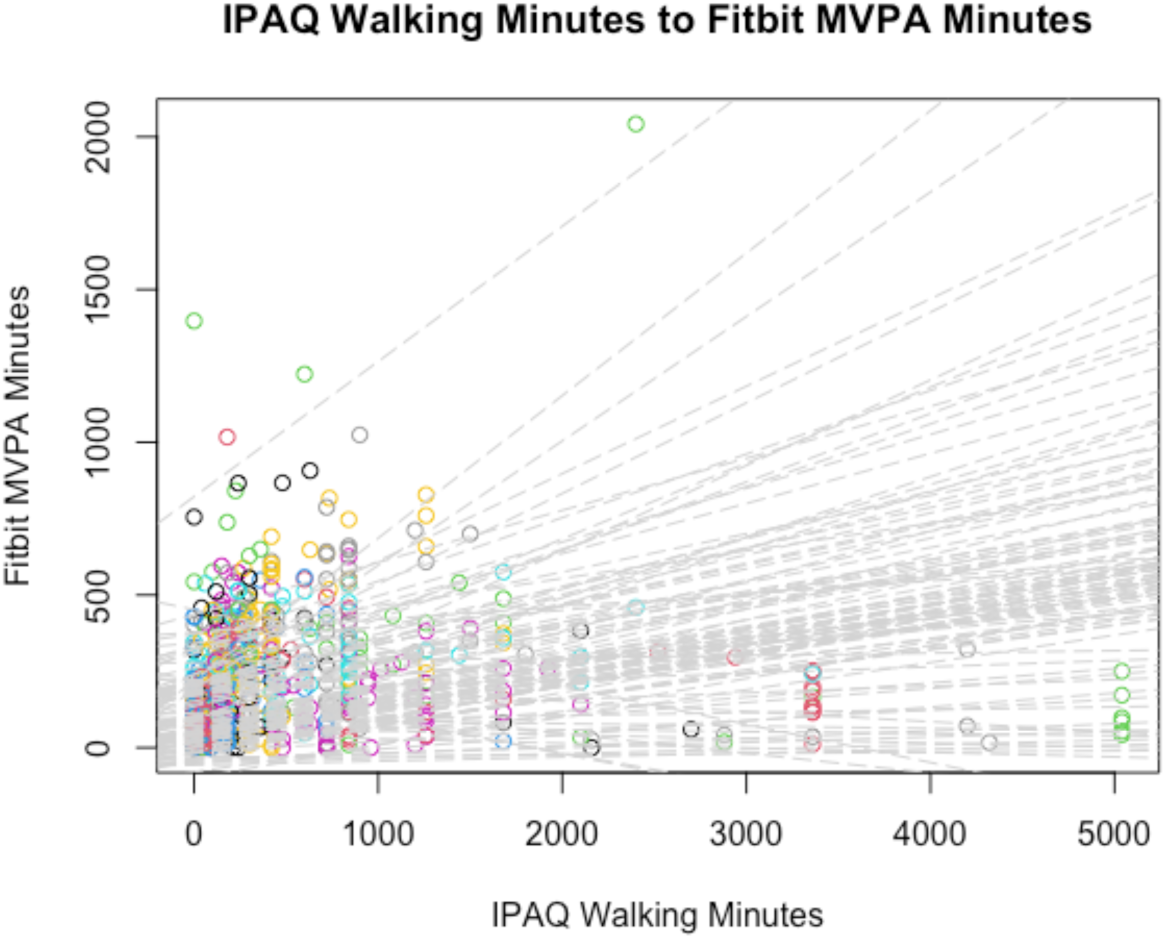
Model-estimated random (i.e., person-level) slopes for the model predicting Fitbit MVPA minutes (y-axis) from IPAQ-based walking minutes. Each participant is represented by one dotted grey line (n=102). Each colored circle represents one person-level week (N=602).

**Figure 3.**
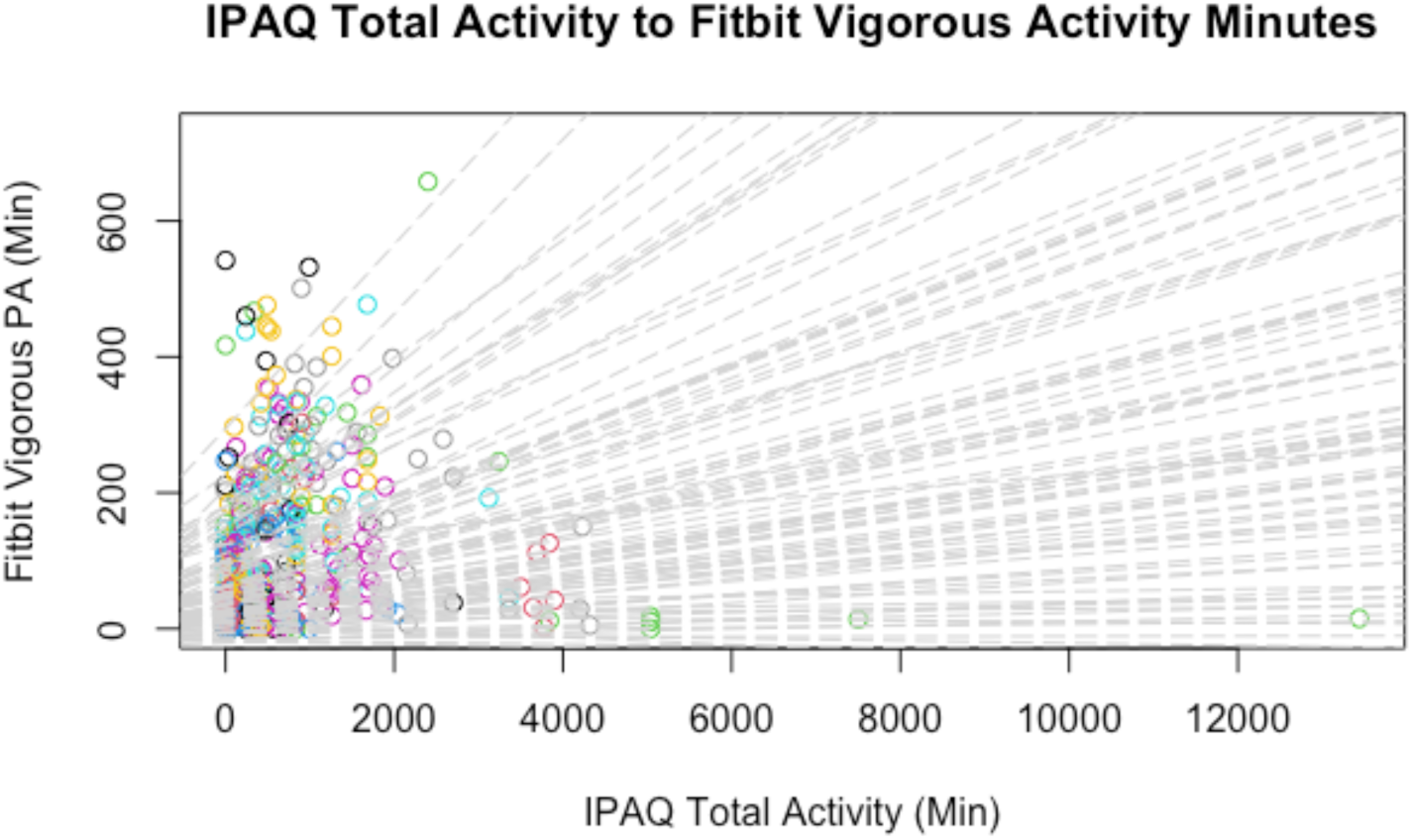
Model-estimated random (i.e., person-level) slopes for the model predicting Fitbit Vigorous PA minutes (y-axis) from IPAQ-based total activity minutes. Each participant is represented by one dotted grey line (n=88). Each colored circle represents one person-level week (N=423).

**Figure 4.**
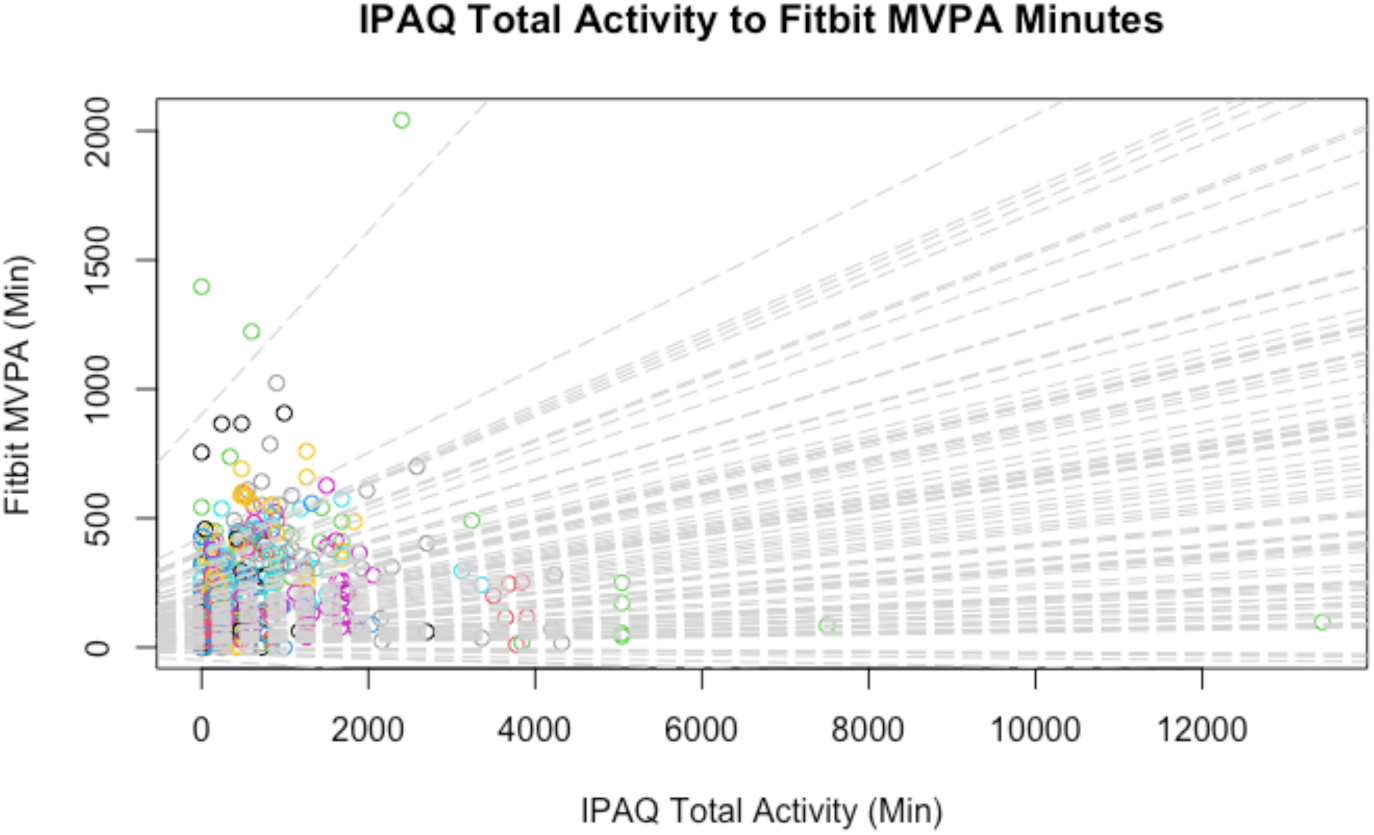
Model-estimated random (i.e., person-level) slopes for the model predicting Fitbit MVPA minutes (y-axis) from IPAQ-based total activity minutes. Each participant is represented by one dotted grey line (n=88). Each colored circle represents one person-level week (N=423).

**Figure 5.**
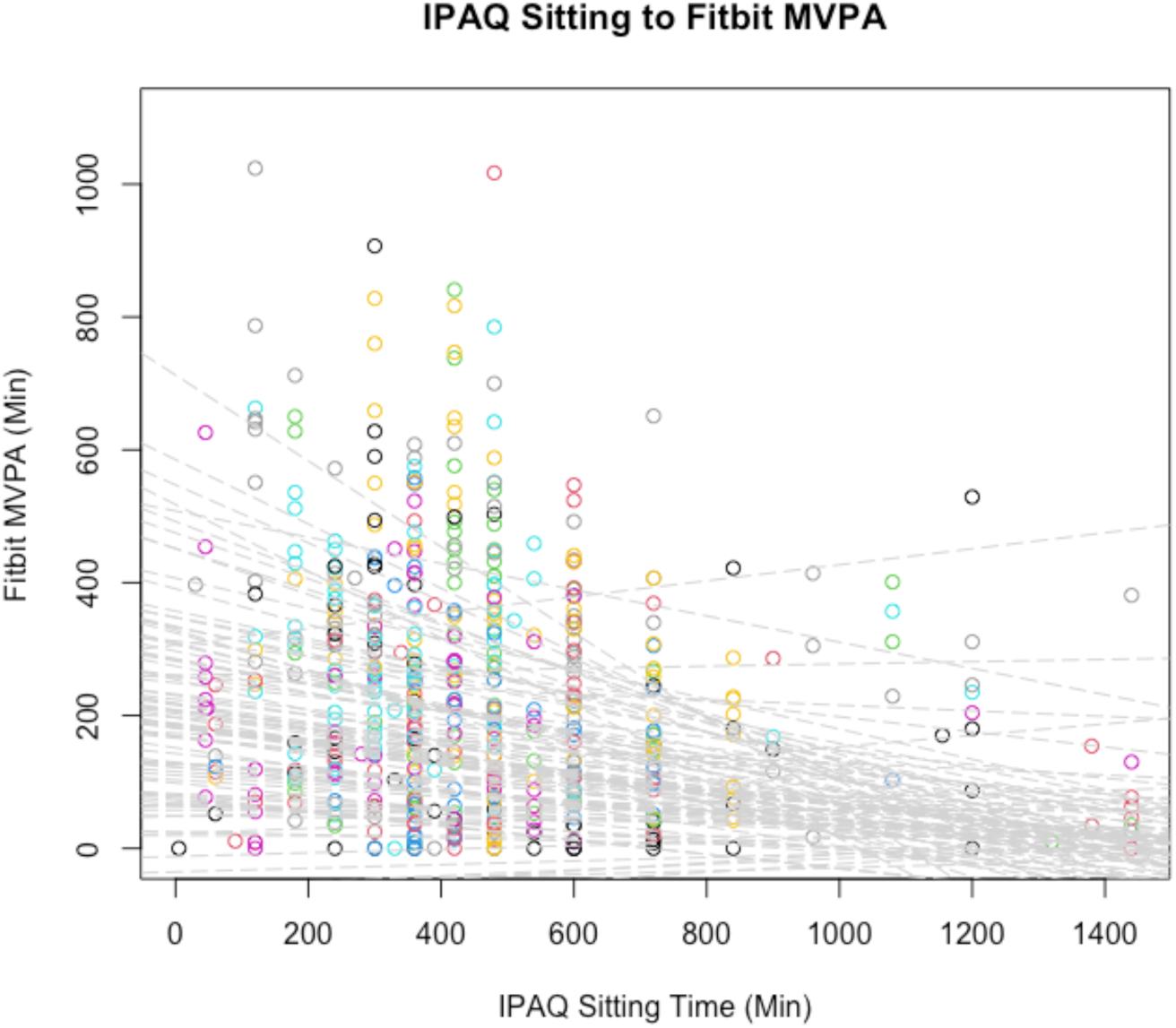
Model-estimated random (i.e., person-level) slopes for the model predicting Fitbit MVPA minutes (y-axis) from IPAQ-based sitting minutes. Each participant is represented by one dotted grey line (n=90). Each colored circle represents one person-level week (N=540).

**Table 3.** Concordance Analysis: Results from 6 separate linear mixed-effects regression models. Point estimates and associated statistics are reported for the fixed effects.

| **Outcome**<br>**(Random slope correlation)** | **Predictors**<br>**(Fixed-Effect Term)** | **B Coefficient (SE)** | **t-value** | **p-value** |
| --- | --- | --- | --- | --- |
| **Fitbit Step Counts**<br>**(Cor = 0.50, p < 0.0001)** | Intercept | 43226.09 (5706.72) | 7.575 | <0.0001 |
|  | IPAQ Walking Min | 10750.15 (2477.63) | 4.339 | 0.0001 |
|  | nweeks | 925.79 (569.45) | 1.626 | 0.106 |
| **Fitbit Light PA Min**<br>**(N/A)** | Intercept | 1050.54 (114.99) | 9.136 | <0.0001 |
|  | IPAQ Walking Min | 66.06 (26.33) | 2.509 | 0.0124 |
|  | nweeks | 30.00 (11.57) | 2.593 | 0.0108 |
| **Fitbit Moderate PA Min**<br>**(Cor = 0.88, p < 0.001)** | Intercept | 87.785 (24.141) | 3.636 | 0.00004 |
|  | IPAQ Walking Min | 36.371 (9.146) | 3.977 | 0.0026 |
|  | nweeks | 3.005 (2.410) | 1.247 | 0.2150 |
| **Fitbit Vigorous PA Min**<br>**(Cor = 0.78, p < 0.001)** | Intercept | 77.749 (27.550) | 2.822 | 0.0056 |
|  | IPAQ Total Activity Min | 29.532 (9.964) | 2.964 | 0.0106 |
|  | nweeks | 4.174 (2.705) | 1.543 | 0.1260 |
| **Fitbit MVPA Min**<br>**(Cor = 0.72, p < 0.001)** | Intercept | 186.269 (43.123) | 4.319 | <0.0001 |
|  | IPAQ Walking Min | 78.721 (22.258) | 3.537 | 0.0014 |
|  | nweeks | 6.872 (4.241) | 1.620 | 0.1079 |
| **Fitbit MVPA Minutes**<br>**(Cor = 0.97, p < 0.001)** | Intercept | 165.196 (50.017) | 3.303 | 0.0012 |
|  | IPAQ Total Activity Min | 57.319 (18.677) | 3.069 | 0.0095 |
|  | nweeks | 6.484 (4.896) | 1.324 | 0.1884 |

The results from the MLMs evaluating divergence (i.e., hypothesized inverse relationship) between IPAQ-reported sitting minutes and the Fitbit-estimated PA scores are provided in Table 4. We report those that indicate statistically significant associations. IPAQ-reported sitting minutes were negatively associated with Fitbit step counts (B = -5222.01, p = 0.0061), MVPA minutes (B = -27.085, p = 0.0136), vigorous PA minutes (B = -13.717, p = 0.0378), moderate PA minutes (B = -11.327, p = 0.0095), and light PA minutes (B = -82.55, p = 0.0167). Reverse-scaling the point estimates, these correspond to decreases in Fitbit step counts by ∼1.36 steps, Fitbit MVPA minutes by 0.0070 minutes, Fitbit vigorous PA minutes by 0.0036 minutes, and Fitbit light PA minutes by 0.021 minutes, for each 1-minute increase in IPAQ-reported sitting time. Finally, none of the random term correlations were significant (See Table 4), though they were all negative as expected.

**Table 4.** Divergence analysis: Results from 5 separate linear mixed-effects regression models. Point estimates and associated statistics are reported for the fixed effects.

| **Model Outcome**<br>**(Random slope correlation)** | **Model Predictor**<br>**(Fixed-Effect Term)** | **B Coefficient (SE)** | **t-value** | **p-value** |
| --- | --- | --- | --- | --- |
| **Fitbit Step Counts**<br>**(Cor = -0.05, p > 0.05)** | Intercept | 39,536.11 (5187.82) | 7.621 | < 0.0001 |
|  | IPAQ Sitting Minutes | -5,222.01 (1535.28) | -3.401 | 0.0061 |
|  | nweeks | 1047.93 (507.98) | 2.063 | 0.0416 |
| **Fitbit Light PA Minutes**<br>**(Cor = 0.09, p = 0.089)** | Intercept | 1105.02 (110.05) | 10.041 | <0.0001 |
|  | IPAQ Sitting Minutes | -82.55 (31.33) | -2.635 | 0.0167 |
|  | nweeks | 20.74 (10.76) | 1.927 | 0.0568 |
| **Fitbit Moderate PA Minutes**<br>**(Cor = -0.97, p = 0.09)** | Intercept | 71.128 (17.795) | 3.997 | 0.0001 |
|  | IPAQ Sitting Minutes | -11.327 (3.753) | -3.018 | 0.0095 |
|  | nweeks | 3.660 (1.730) | 2.116 | 0.0369 |
| **Fitbit Vigorous PA Minutes**<br>**(Cor = -0.48, p = 0.235)** | Intercept | 73.167 (24.677) | 2.295 | 0.0036 |
|  | IPAQ Sitting Minutes | -13.717 (6.026) | -2.276 | 0.0378 |
|  | nweeks | 5.692 (2.410) | 2.362 | 0.0199 |
| **Fitbit MVPA Minutes**<br>**(Cor = -0.61, p = 0.091)** | Intercept | 145.593 (38.493) | 3.782 | 0.0002 |
|  | IPAQ Sitting Minutes | -27.085 (9.250) | -2.928 | 0.0136 |
|  | nweeks | 9.178 (3.751) | 2.447 | 0.0162 |

### Moderation by habitual MVPA

Results of the mixed effects model estimating the moderator effect of average weekly (i.e., habitual) MVPA are provided in Table 5. The point estimate for the interaction term indicated that overall, habitual MVPA positively moderates the relationships between IPAQ total activity minutes and Fitbit vigorous PA minutes (B = 20.214, p < 0.0001), IPAQ walking minutes and Fitbit step counts (B = 5045.209, p < 0.0001), and IPAQ walking minutes and Fitbit moderate PA minutes (B = 34.080, p < 0.0001). Reverse-scaling, every 1-minute increase in IPAQ-reported total activity was associated with an increase in Fitbit vigorous PA minutes by 0.018 minutes, when controlling for habitual MVPA levels. Similarly, for every 1-minute increase in IPAQ-reported walking, Fitbit step counts increase by ∼4.81 steps, Fitbit light PA minutes increase by 0.053 minutes, and Fitbit moderate PA minutes increase by 0.021 minutes, when controlling for habitual MVPA. The models controlled for habitual physical activity measuring the relationship of Fitbit vigorous, moderate, and sedentary minutes to their corresponding IPAQ variable were not significant (p > 0.05).

**Table 5.** Results from 4 separate linear mixed-effects regression models that are estimating IPAQ self-reported activity on different types of Fitbit data (step count, lightly active minutes, and vigorous activity minutes minutes).

| **Model Outcome** | **Model Predictors (Fixed effect term)** | **B Coefficient (SE)** | **t-value** | **p-value** |
| --- | --- | --- | --- | --- |
| **Fitbit Step Counts** | Intercept | 51317.551 (3401.414) | 15.087 | < 0.0001 |
|  | IPAQ Walking Min | 5785.888 (977.868) | 5.917 | < 0.0001 |
|  | Habitual MVPA | 20043.456 (1231.025) | 16.282 | < 0.0001 |
|  | IPAQ Walking Min * Habitual MVPA | 5045.209 (893.586) | 5.646 | < 0.0001 |
|  | nweeks | 0.166 (330.233) | 0.001 | 1 |
| **Fitbit Light PA Min** | Intercept | 1174.25 (107.53) | 10.921 | < 0.0001 |
|  | IPAQ Walking Min | 63.56 (25.62) | 2.481 | 0.0134 |
|  | Habitual MVPA | 201.67 (42.13) | 4.787 | < 0.0001 |
|  | IPAQ Walking Min * Habitual MVPA | 19.59 (21.26) | 0.921 | 0.3573 |
|  | nweeks | 17.74 (10.75) | 1.650 | 0.1019 |
| **Fitbit Moderate PA Min** | Intercept | 123.322 (15.149) | 8.140 | < 0.0001 |
|  | IPAQ Walking Min | 25.507 (4.231) | 6.029 | < 0.0001 |
|  | Habitual MVPA | 92.991 (5.581) | 16.661 | < 0.0001 |
|  | IPAQ Walking Min * Habitual MVPA | 34.080 (3.785) | 9.003 | < 0.0001 |
|  | nweeks | -0.737 (1.481) | -0.498 | 0.62 |
| **Fitbit Vigorous PA Min** | Intercept | 131.322 (14.873) | 8.829 | < 0.0001 |
|  | IPAQ Walking Min | 9.167 (4.516) | 2.030 | 0.043 |
|  | Habitual MVPA | 87.267 (5.131) | 17.008 | < 0.0001 |
|  | IPAQ Walking Min * Habitual MVPA | 18.133 (4.364) | 4.155 | < 0.0001 |
|  | nweeks | -0.408 (1.416) | -0.288 | 0.773 |
| **Fitbit Vigorous PA Min** | Intercept | 122.115 (17.539) | 6.962 | < 0.0001 |
|  | IPAQ Total Activity Min | 15.446 (5.119) | 3.017 | 0.0027 |
|  | Habitual MVPA | 84.555 (5.933) | 14.251 | < 0.0001 |
|  | IPAQ Total Activity Min * Habitual MVPA | 20.214 (4.945) | 4.088 | < 0.0001 |
|  | nweeks | -0.062 (1.670) | -0.038 | 0.9700 |

## Discussion

This study aimed to evaluate scores from the IPAQ against objectively estimated PA scores from Fitbit devices in women with CPPDs. Accurate, comprehensive PA measurement whilst minimizing participant burden and resource requirements is important for managing symptoms and improving quality of life, however, the utility and validity of self-report PA tools like the IPAQ remains underexplored in CPPDs.^1^ The strong associations between several IPAQ- and Fitbit-based scores suggest that the IPAQ can provide reasonable estimates of some activities (e.g., walking) in women with CPPDs. We further report systematic variations across individuals in PA reporting, supported by variance over time and effect of habitual MVPA levels. To our knowledge, this is the first study to evaluate the IPAQ in individuals with CPPDs and through longitudinal data.

The results from the ICC analyses provide additional insights into the temporal agreement patterns between the two methods. The walking minutes/steps domain demonstrated the highest consistency for both the IPAQ and the Fitbit (i.e., R = 0.766 and R=0.601, respectively), followed by total PA (i.e., R=0.646 and R=0.470, respectively). It is possible that walking behavior is easier to recall due to its routine and identifiable nature. Previous studies report a wide range of ICCs for IPAQ-based walking minutes,^19,39–43^ though the data collection period in those studies were 2 weeks or shorter. Next, we observed that IPAQ-reported MVPA, vigorous, and moderate PA minutes had lower consistency compared to their Fitbit counterparts (Table 2). This may indicate difficulty in accurately categorizing higher intensity PA through self-report, a finding in line with other similar studies.^44^ The consistently higher ICC2a values with the inclusion of the interaction term suggest systematic patterns in individual PA reporting relative to Fitbit estimates, most evident in total PA and sedentary behavior domains. Walking minutes/steps demonstrated the most consistent agreement (ICC2r = ICC2a = 0.402). The low ICC2r values for vigorous and moderate PA with improvements in ICC2a when accounting for interaction indicate poor absolute agreement but consistent relative patterns within individuals. Finally, sitting minutes demonstrated moderate to strong temporal stability for both IPAQ (R = 0.595) and Fitbit (R = 0.810) measures, yet indicated poor inter-method agreement (ICC2r = 0.101). This demonstrates that while the measurements remain relatively consistent over time, there are substantial differences in how this behavior is captured or defined between the two methods. This is expected given the IPAQ and the Fitbit measure somewhat different constructs (self-reported sitting vs. any movement that falls below the threshold for light intensity). As such, exact conceptual equivalence cannot be assumed.^45^ Taken together, these results suggest that IPAQ and Fitbit may be more suitable for tracking individual-level changes over time, particularly based on walking behavior and total PA. They also highlight the importance of considering person-specific measurement properties in PA assessment, a key principle in measurement science.^46^

The results of the validity evaluation through MLMs indicated that IPAQ-reported walking minutes were positively associated with numerous Fitbit-measured outcomes including step count, MVPA, moderate activity minutes, and light PA minutes. Similarly, IPAQ total activity minutes indicated good concordance with vigorous PA and MVPA measured by Fitbit. These findings suggest that the IPAQ item on walking minutes might be the most efficient for retrospective PA assessment in this population, whereas total PA might be useful in estimating higher-intensity activities. As such, these two items may be possible replacements in instances where resource constraints limit the use of accelerometer devices,^9^ or when wearing continuous activity monitors is not feasible. A study validating the IPAQ in visually impaired adults^47^ similarly reported that IPAQ walking minutes were predictive of steps, light PA and total PA, but not MVPA. In contrast, another study^42^ in healthy college students (74% female) demonstrated that whilst IPAQ-reported walking minutes did not correlate with any of the Fitbit-based PA outcome scores, moderate- and vigorous intensity PA minutes were associated with their Fitbit-based counterparts as well as step counts. The discrepant findings could be due to the shorter study period (1 week), use of ActiGraph devices, or differences in sample demographics.^48^ Nevertheless, these results underscore the importance of considering population-specific factors when interpreting inferences made from self-reported PA data or choice of PA domain when designing interventions^4,6^ (e.g., MVPA for targeting pain management,^49^ step counts for targeting depressive symptoms or physical function).^50,51^

We further report individual variability in the relationship between IPAQ-reported and Fitbit-measured PA outcomes, based on the random slope terms in the MLMs. These effects were significant for step counts, MVPA, vigorous and moderate intensity PA (Table 3), and point to individual differences in PA reporting among women with CPPDs. The strong correlations between the random intercepts and slopes corroborate the non-significant point estimates observed for the fixed-effect terms (i.e., average group effect) in these models. That is, the magnitude of variance between groups (i.e., level - 2 random effects) will in principle be inverse to the magnitude of a fixed-effect term (i.e., level -1 predictor). Accordingly, the generalizability of these results appears strongest for walking-based assessments. In contrast, IPAQ-reported sitting minutes were not significantly associated with Fitbit-estimated sedentary minutes. They were, however; significantly and inversely associated with other domain scores, providing support for divergence between these constructs. Moreover, these associations were homogenous across the sample based on the non-significant random slope term. Our findings on the low concordance in sedentary time estimates align with those from several other studies using a variety of devices (e.g.,GT1M ActiGraph accelerometer^52,53^, SenseWear Pro Armband; SWA^19^, Xiaomi Mi Band 2^14^). Collectively, these results suggest that sedentary time is relatively more challenging to accurately recall and report regardless of population demographics.

Finally, our analyses identified habitual MVPA levels as a moderator of the relationship between IPAQ walking minutes and Fitbit step counts and Fitbit moderate PA minutes, as well as between IPAQ total activity minutes and Fitbit vigorous PA minutes (See Table 5). It is possible that individuals who are more active have better awareness and recall of their activity patterns, based on the stronger associations between their IPAQ-reported and Fitbit-estimated PA scores. On the other hand, the interaction term was not significant for predicting Fitbit-based light PA. This pattern of significance follows that of the random slope terms (Table 3). One explanation is that the between-individual variance in those models is explained away once habitual MVPA is included as an interaction term (Table 5). This would then suggest that the unobserved between-individual differences were largely due to habitual MVPA differences and/or closely linked factors. For example, habitual MVPA was demonstrated to predict weight change, weight compensation, and changes in energy intake.^54^ Taken together, these results demonstrate that an individual’s typical activity level influences their reporting accuracy, and interpretation of IPAQ-reported PA scores are more generalizable to individuals with similar habitual activity levels within the CPPD population.

## Limitations

We acknowledge several limitations of this study. First, the study sample was relatively homogenous in demographic factors, including education and employment status. These demographic characteristics may influence PA levels and self-reporting, potentially limiting the generalizability of the findings. We did not include these as additional moderators in our analyses because of the relative homogeneity in the sample and they were beyond the scope of our *a priori* objectives. Future studies may benefit from additionally evaluating these factors based on evidence for their influence on PA behavior and self-reporting.^55,56^ The use of Fitbit devices also poses limitations, including potential under- or overestimation of various PA parameters.^57–59^ They may not accurately capture certain types of activities, such as cycling, swimming,^60^ weightlifting or stationary exercises.^61^ In contrast to prior studies using 1-2 weeks of data, the longer study period herein enables a more robust and comprehensive analyses. Nevertheless, some participants in our sample had fewer weeks of data, prompting us to adjust for this variability in our regression analyses. The effect of number of weeks were not significant for most of the models, however; the magnitude of the point estimates could increase with more data points, especially for those with higher variability (i.e., lower ICCs).

## Conclusion

In conclusion, this study provides insights into the strengths and limitations of using the IPAQ for assessing PA in women with CPPDs. The strong associations between IPAQ walking minutes and various Fitbit-measured PA scores suggest that self-reported walking might be a more comprehensive indicator of overall PA than previously thought in this population. Our findings further point to individual differences in PA reporting, which underscores the need for caution in generalizing results across all individuals with CPPDs and considering systematic differences in reporting across individuals. Similarly, considering habitual activity levels when interpreting self-reported PA data is warranted, as more active individuals demonstrated greater concordance in their self-reports. The IPAQ can be a useful complementary tool for PA measurement in clinical and research settings, especially when resources limit the use of more expensive accelerometer devices.

## Funding Statement

This study was supported by the Eunice Kennedy Shriver National Institute Of Child Health & Human Development of the National Institutes of Health under Award Number R01HD108263 (PI=Ensari). The content is solely the responsibility of the authors and does not necessarily represent the official views of the National Institutes of Health.

## Data Availability Statement

The data used in this study can be made available after the active grant period is over upon reasonable request to the corresponding author.

## Notes

### Competing Interest Statement

The authors have declared no competing interest.

### Author Declarations

The Institutional Review Board (IRB) of the Icahn School of Medicine at Mount Sinai (ISMMS) gave ethical approval for this work.

